# The iCARE feasibility non-experimental design study: An integrated collection of education modules for fall and fracture prevention for healthcare providers in long term care

**DOI:** 10.1101/2024.03.22.24304705

**Authors:** Isabel B. Rodrigues, George Ioannidis, Lauren Kane, Loretta M. Hillier, Caitlin McArthur, Jonathan Adachi, Lehana Thabane, George Heckman, Jayna Holroyd-Leduc, Susan Jaglal, Sharon Kaasalainen, Sharon Straus, Momina Abbas, Jean-Eric Tarride, Sharon Marr, John Hirdes, Arthur N. Lau, Andrew Costa, Alexandra Papaioannou

## Abstract

Falls and hip fractures are a major health concern among older adults in long term care (LTC) with almost 50% of residents experiencing a fall annually. Hip fractures are one of the most important and frequent fall-related injuries in LTC. The purpose of this study was to determine the feasibility (recruitment rate and adaptations) of implementing the PREVENT (Person-centred Routine Fracture PreEVENTion) model in practice, with a subobjective to understand facilitators and barriers. The model includes a multifactorial intervention on diet, exercise, environmental adaptations, hip protectors, medications (including calcium and vitamin D), and medication reviews to treat residents at high risk of fracture. Our secondary outcomes aimed to assess change in knowledge uptake of the guidelines among healthcare providers and in the proportion of fracture prevention prescriptions post-intervention. We conducted a mixed-methods non-experimental design study in three LTC homes across southern Ontario. A local champion was selected to guide the implementation. We reported recruitment rates using descriptive statistics and adaptations using content analysis. We reported changes in knowledge uptake using the paired sample t-test and the percentage of osteoporosis medications prescriptions using absolute change. Within five months, we recruited three LTC homes. We required two months to identify and train the local champion over three 1.5-hour train-the-trainer sessions, and the champion required three months to deliver the intervention to the healthcare team. We identified several facilitators, barriers, and adaptations. Benefits of the model include easy access to the Fracture Risk Scale, clear and succinct educational material catered to each healthcare professional, and an accredited educational module for physicians and nurses. Challenges included misperceptions between the differences in fall and fracture prevention strategies, fear of perceived side effects associated with fracture prevention medications, and time barriers with completing the audit report. Our study did not increase knowledge uptake of the guidelines, but there was an increase in the proportion of osteoporosis medication post-intervention.

## INTRODUCTION

Falls and hip fractures are a major health concern among older adults (1). An estimated one in three individuals aged 65 years and older fall each year, and almost half of individuals living in long term care (LTC) fall annually (2). Hip fractures in residents living in LTC are devastating and are likely to increase with current demographic trends in aging (3,4). Each year, almost 50% of residents in LTC facilities fall at least once, and 40% of residents fall twice or more (3,5). Of those LTC residents that fall, 10% to 25% of falls are associated with serious injuries requiring medical treatment with 2% to 6% resulting in fractures of the hip, wrist, or vertebra (6–8). Hip fractures are particularly life altering as both mobility and the capacity to live independently are affected (3,6–8). Moreover, about half of the residents in LTC who experience a hip fracture may die within the next year or develop total dependence within six months of their hip fracture (3,6–8). Thus, interventions to reduce falls and fractures in LTC are important to decrease morbidity and mortality.

In 2015, the Canadian recommendations for preventing fractures in LTC were released to facilitate an evidence-based decision-making process (9). Indeed, guidelines have the capacity to promote high quality practice informed by evidence, enable appropriate resource allocation, and advance research by identifying knowledge gaps (10); however, the existence of guidelines alone is not enough to change practice (10,11). The lack of guideline uptake in practice is evident with the ongoing high morbidity and mortality associated with fractures in LTC (12–14). The major gap is the limited evidence on effective knowledge translation strategies to prevent falls and low trauma fractures in residents who are at high risk of fractures (12,15,16). Active strategies that utilize an integrated knowledge translation approach may help to uptake guidelines into practice (12). Current methods to uptake guidelines involve co-developing models with end-users and stakeholders and piloting their implementation under real-world conditions (17).

PREVENT (<u>P</u>erson-centred <u>R</u>outine Fracture Pre<u>EVENT</u>ion) is an educational outreach model for delivering education on fall and fracture prevention to healthcare providers working in LTC. The model includes a multifactorial intervention on diet, exercise, environmental adaptations to reduce falls, hip protectors, medication (including calcium and vitamin D), and medication reviews. The model, herein known as PREVENT, aligns with the 2015 Canadian evidence-based recommendation (9) and with recommendations from Australia and USA (18). PREVENT utilizes a multifactorial knowledge mobilization strategy that engages the entire multi-disciplinary LTC team (e.g., physicians, nurses, physiotherapists, dietitians, personal support workers) and builds automated identification of high-risk fractures into routine care processes.

PREVENT may have several benefits including reducing falls and fractures and improving quality of life of residents in LTC. Before a trial can be implemented to determine the effectiveness of PREVENT in practice, we need to determine the feasibility of implementing it under real-world conditions. The purpose of the iCare (integrated <u>c</u>ollection of education modules for f<u>a</u>ll and fracture pr<u>e</u>vention) study was to determine the feasibility of implementing PREVENT to healthcare providers in LTC. Our secondary outcomes were to determine if PREVENT improved knowledge uptake of the Canadian recommendations (9) and increased the proportion of osteoporosis medication prescriptions post-intervention. We selected the proportion of change in osteoporosis medications since these medications currently have the highest certainty of evidence to reduce the risk and the rate of fractures (19,20) and are the easiest outcome to measure in practice.

## METHODS

### Study design

We conducted a pre-post non-experimental design study in three LTC homes across Ontario. We conducted our study in accordance with the STROBE 2007 guidelines (21) and TIDieR 2014 checklist (22). We registered our study on clinicaltrials.gov on July 6, 2022 (NCT05445336); we updated the registry on December 21, 2022. We received ethics approval from the Hamilton Integrated Research Ethics Board (Project ID 14463).

### Study setting

Between January to June 2022, we sent emails to five large LTC corporate offices detailing the iCARE study. Each corporation owned between three to 30 homes across Ontario. Two LTC corporations were willing to participate in the feasibility study. We recruited one for-profit and two not-for profit LTC homes, herein known as Home A (120 beds), Home B (425 beds), and Home C (240 beds). All three LTC homes are in southern Ontario, Canada in large metropolitan cities.

### Knowledge translation model

PREVENT is an organizational-level, service provision approach that embeds falls and fracture education and management strategies in LTC. The model utilizes several intervention functions and behaviour change techniques based on the Behaviour Change Wheel (Table 1) (23,24). The specific PREVENT components are detailed in S1.

**Table 1:** Intervention functions and behaviour change techniques used in PREVENT.

| **Intervention function from Behaviour Change Wheel** | **Definition of intervention function** | **Behaviour change technique** | **Behaviour change technique example** |
| --- | --- | --- | --- |
| Education | Improving knowledge and understanding of roles and responsibilities for healthcare professionals responsible for service provision and improving knowledge through a community of practice (24) | • Feedback on behaviour<br>• Prompts/cues<br>• Self-monitoring of behaviour | • Audit and Feedback Report |
| Training | Providing key professionals with core skills in management and leadership (24) | • Demonstration of the behaviour<br>• Instruction on how to perform a behaviour | • Train the trainer<br>• Educational videos |
| Modelling | Discussion of identified successful strategies for change within network (24) | • Demonstration of the behaviour | • Train the trainer<br>• Educational videos<br>• Training manual<br>• Fracture and Fall Prevention Guide |

PREVENT was designed to be integrated into regularly scheduled healthcare sessions (e.g., Professional Advisory Committee, Falls Committee), which usually occur quarterly in LTC. We identified a local champion to help guide the implementation of the model and continue to promote best practices (25). The standardized model includes: 1) train-the-trainer sessions with the local champion (26); 2) a multidisciplinary educational session with fracture prevention care recommendations and real-world case studies (27); 3) orientation to the Fracture Prevention Toolkit developed by the lead researcher (IBR) that is provided to each home with the Fracture Risk Scale (FRS) which identifies residents at high risk of fracture (score between 4 to 8) and low risk of fracture (score between 1 to 3) (28), templates, standard order sets, and quick reference guide to the 2015 Canadian LTC fracture prevention guidelines (29); 4) an audit and feedback report developed by the local champion for physicians and healthcare providers regarding the number of residents at high versus low risk of fracture, number of residents who have experienced a fragility fracture in the last six months, and the number of high risk of fracture residents on osteoporosis medications (30); and 5) implementation intentions that includes an implementation proposal template, care planning template, and Process Indicator Checklist (home-level policies that support and sustain fracture prevention best practices such as training and education of front-line staff including personal support workers) (31). We utilized a three-step process to implement PREVENT. In step one (train-the-trainer), the research team worked with each home to identify a local champion. We sought champions who the home’s management team considered trustworthy and influential, acted as a role model for behaviour change, supported and legitimized the work, provided a mechanism of communication between the research team and healthcare providers in LTC, and served to sustain gains in the long term (25). The lead investigator (IBR) met with each local champion using an online platform (i.e., Zoom) and in-person at the LTC home to review how to deliver PREVENT, which included reviewing how to use the FRS to identify residents at high and low risk of fracture (28), a training manual (see S2), Fracture and Fall Prevention Guide (see S3), educational module, and other point of care tools. Our processes were guided by the Getting to Outcomes Framework, which includes a 10-step program for implementing, evaluating, and continuously improving prevention programs (32). In step two, the local champion adapted the educational meeting to the LTC home and developed the audit and feedback report. During the third step, the local champion presented the adapted PREVENT material during the leadership team meeting which included physicians, nurse practitioners, registered nurses, dietitians, physiotherapists, kinesiologists, personal support workers, and interRAI coordinators. InterRAI coordinators are part of the interRAI network and are responsible for collecting health outcomes in LTC (https://interrai.org/about-interrai/). During the meeting, the leadership team also brainstormed and implemented intentions and care plans to identify and treat residents at high risk of fracture. LTC homes were remunerated for their time and participation in the iCARE study.

### Intervention timeline

|  | Intervention Timeline |
| --- | --- |
| Month 1 and Month 2 | 1. LTC home recruitment<br>2. Identify local champion |
| Month 3 | 1. Train-the-trainer to prepare audit and feedback report and adapt/deliver the educational module |
| Month 4 | **Interactive Session 1**<br>1. Local champion leads education module with the leadership team<br>2. Audit and feedback report review<br>3. Develop action and care plan |
| Months 4 to Month 6 | 1. LTC home implements action and care plan |
| Month 7 | **Interactive Session 2**<br>1. Focus group with local champion and leadership team to understand barriers, facilitators, and adaptations<br>2. One-on-one interview with local champion to understand barriers, facilitators, and adaptations |

### Inclusion/exclusion criteria

As this is a pragmatic study, our inclusion and exclusion criteria were broad and generalizable to represent real world practice. Our organizational level inclusion criterion was to include homes with a minimum of 70 beds, which is the sample size for our larger clinical trial (i.e., PREVENT trial).

### Outcomes

#### Primary outcome

The primary outcome was feasibility of implementation defined by the number of LTC homes recruited within two months, length of time for the local champion to deliver the PREVENT program, and the length of time it took the local champion to develop the audit and feedback report (Table 2) (33). The lead investigator (IBR) developed a checklist prior to the start of the study to assess fidelity; our method to develop the checklist was defined using a five-step guide proposed by Michie and colleagues (34,35). The fidelity checklist assessed domains related to preparing the educational session (e.g., audit reporting, scheduling a date and time for the leadership team to meet), presenting the educational session (e.g., reviewing the FRS and guidelines, developing, and reviewing the audit and feedback report), and leading discussions on the action and care plan processes. The fidelity checklist included 23 items, and each item was rated from 0 (action was not completed), 0.25 (action was 25% completed), 0.50 (50% completed), 0.75, and 1.00. A total score of 85% (20/23) was considered good fidelity, moderate between 50% to 84%, and poor <50% (34,35). The fidelity checklist was completed by the lead investigator (IBR) during the education session. To understand adaptations after implementing the intervention, we held focus groups with the local champion and the leadership team. Focus groups were recorded and transcribed verbatim. We also held informal one-on-one interviews using open-ended questions with each local champion to identify adaptations to the model; these interviews were held three months after the educational session and led by the lead researcher. The one-on-one interview with the local champion was not recorded, but detailed field notes were taken by the research coordinator (LK).

**Table 2:** Feasibility outcomes and criteria for success (33)

| Feasibility outcomes | Criteria for success |
| --- | --- |
| Recruitment rate (organizational level – length of time to recruit three LTC homes) | Recruit three LTC homes in Ontario within 2 months |
| Recruitment rate (participant level - length of time to identify and train local champion) | Two months to identify, meet, and train local champion |
| Determine challenges to deliver the PREVENT model and program | ≥ 80% on the fidelity checklist<br>Identify facilitators of and barriers to implementation during the focus group |
| Determine challenges in linking the data for the audit and feedback report | Identify facilitators of and barriers to developing the audit and feedback report |

#### Secondary outcomes

Our secondary outcomes were to determine if there was a change in knowledge uptake and in the proportion of osteoporosis medications before and after the intervention. We assessed change in knowledge uptake using a pre-post test. First, we developed a case study of a typical resident living in LTC. Next, healthcare providers were asked to complete a series of multiple-choice questions about identifying the resident’s FRS, prescribing medications, supplements, and exercise, conducting an environmental hazard scan, and using hip protectors. Immediately after the education session, we asked healthcare providers to repeat the multiple-choice questionnaire on the same case study. We collected data on information on osteoporosis medications at baseline (six months prior to implementing the intervention) and post-intervention (three to four months after the intervention). We collected information on the number of residents at high risk of fracture or low risk of fracture using the FRS, the number of residents on an osteoporosis medication (e.g., bisphosphonates, denosumab), and the number of osteoporotic fractures at baseline and post-intervention. We defined fractures as any major osteoporotic fracture of the hip, pelvis, vertebrae, or distal radius (36,37). The local champion pulled data on FRS scores, fractures, and medications directly from all residents’ charts in the electronic medical record or the Ontario Drug Benefit database. A list of osteoporosis medications is provided in S4 table.

### Analysis

We reported our recruitment rates and fidelity scores using descriptive statistics as a total score or value. We reported the results of the focus groups using content analysis (descriptive qualitative analysis); two reviewers (IBR and LK or LH) coded the data independently using NVivo, version 14 (QSR International Pty Ltd, Doncaster, VIC, Australia) (38). The reviewers then met to discuss and compare the codes and group the codes into categories (38). Disagreements were resolved by the third reviewer (38). We collected demographic characteristics of the local champion and leadership team, which were reported using the number of individuals and a percentage. To assess change in knowledge uptake, we conducted a paired sample t-test (two-sided p). To report changes in osteoporosis medications, we conducted two data pulls at baseline and post-intervention; we included residents who “moved-in” (i.e., moved into the home during the post-intervention phase) and “moved-out” (i.e., passed away during the post-intervention phase) in the analysis. We reported the results using absolute change. Quantitative results were analyzed using IBM SPSS Statistics for Windows, version 28 (Armonk, NY: IBM Corp).

## Results

### Feasibility

We recruited three LTC homes between October 2022 and February 2023; our recruitment process was affected by residual COVID-19 complications. After we identified a home, the home’s administrative team took approximately a month to identify a potential local champion. Two of the local champions were nurse practitioners, and the third champion was a kinesiologist. Subsequently, our research team met with the local champion to complete the train-the-trainer sessions. Our research team completed two, one-hour train-the-trainer sessions with Home A; however, based on feedback from Home A, we added a third train-the-trainer session. The local champion led the third session where they practiced delivering the educational module to the research team. Home B and C received three sessions and each session was 1.5 hours in length. After the train-the-trainer sessions, the local champion required at least two months to review the educational materials, adapt the material to their home, and develop the audit and feedback report. All leadership team meetings were held in-person at a convenient time at their respective LTC home. The characteristics of the local champion and leadership team are presented in Table 3. PREVENT required at least 10 months per home to recruit, train, and implement.

**Table 3:** Local champion and leadership team characteristics (n = 3)

|  | **Home A**<br>**(k=10)** | **Home B**<br>**(k=19)** | **Home C**<br>**(k=11)** |
| --- | --- | --- | --- |
| **Local champion** |  |  |  |
| Highest level of education | College | University/post-graduate | University/post-graduate |
| Fidelity score (total score 23 points) | 19.75/23 (86%) | 20.25/23 (88%) | 20.00/23 (87%) |
| **Leadership team** |  |  |  |
| Gender (female), n (%) | 8 (80%) | 8 (42%) | 10 (91%) |
| Level of Education, n (%) |  |  |  |
| College | 5 (50%) | 4 (21%) | 1 (9%) |
| University | 5 (50%) | 8 (42%) | 9 (82%) |
| Prefer not to answer | 0 (0%) | 7 (37%) | 1 (9%) |
| Role in LTC home, n(%) |  |  |  |
| Physician | 0 (0%) | 4 (21%) | 1 (9%) |
| Pharmacist | 0 (0%) | 1 (5%) | 1 (9%) |
| Administrator | 1 (10%) | 1 (5%) | 0 (0%) |
| Director of care/Nursing director | 1 (10%) | 2 (11%) | 1 (9%) |
| Neighbourhood coordinator | 2 (20%) | 4 (21%) | 2 (18%) |
| Dietitian | 0 (0%) | 1 (5%) | 1 (9%) |
| Nurse practitioner | 0 (0%) | 1 (5%) | 2 (18%) |
| Nursing student | 1 (10%) | 0 (0%) | 0 (0%) |
| Registered nurse | 1 (10%) | 0 (0%) | 0 (0%) |
| Rehabilitation specialist/Physiotherapist | 2 (20%) | 2 (11%) | 2 (18%) |
| Personal support worker | 2 (20%) | 3 (16%) | 1 (9%) |
| Length of time worked within current profession in LTC, n (%) |  |  |  |
| < 1 year | 2 (20%) | 1 (5%) | 1 (9%) |
| 1 to 2 years | 0 (0%) | 2 (11%) | 1 (9%) |
| 3 to 5 years | 2 (20%) | 1 (5%) | 1 (9%) |
| > 5 years | 5 (50%) | 8 (42%) | 7 (64%) |
| Prefer not to answer | 1 (10%) | 7 (37%) | 1 (9%) |
| Length of time providing care to residents at risk of falls and fractures, n (%) |  |  |  |
| < 1 year | 2 (20%) | 1 (5%) | 1 (9%) |
| 1 to 3 years | 1 (10%) | 0 (0%) | 1 (9%) |
| 3 to 5 years | 6 (60%) | 10(53%) | 8 (73%) |
| > 5 years | 1 (10%) | 1 (5%) | 0 (0%) |
| Prefer not to answer | 0 (0%) | 7 (37%) | 1 (9%) |

We had a diverse group of healthcare professionals on the leadership team (Table 3). In Home A, the local champion recruited ten healthcare professionals to the leadership team, in Home B, 19 healthcare professionals, and in Home C, 12 healthcare professionals. The attendance rate during the leadership team for Home A was 90% (9/10), Home B, 84% (16/19), and Home C, 90% (10/11). Fidelity scores ranged from moderate to good (Table 3).

### Challenges, successes, and adaptations

During the focus group, each home identified similar barriers to and facilitators of implementing PREVENT in practice.

#### Challenges

Challenges to implementing PREVENT included having an unclear sustainability strategy, misperceptions between fall and fracture prevention strategies, fear of prescribing osteoporosis medications, and time barriers with completing the audit report. During the focus group, the local champion expressed that it was unclear who would sustain the role of implementing PREVENT after the research team had left. The local champion cited lack of time and other commitments as a barrier to sustaining the model; all three champions noted that a major barrier was dedicating time to the audit and feedback report. To manually extract the data and link the variables to each resident for the audit report, the local champion required at least one full day per week for five weeks (i.e., 40 hours total). Moreover, several leadership members were unsure of the implementation plan after identifying a resident at high risk of fracture:

> “I found myself often sometimes ignoring the prompts for FRS <Fracture Risk Scale> unless there was a specific reason. Cold calling families to say your fracture risk scale is high, and that’s the only reason I’m calling is weird.” [Home B]

Several healthcare professionals on the leadership team felt the strategy to prevent fractures (i.e., PREVENT) was akin to their current fall prevention strategy. It was perceived that if a healthcare professional could prevent a fall, it would translate to preventing a fracture.

> “*I think a lot of homes already are looking at all the PREVENT aspects, it’s just not known as PREVENT or they’re not grouping them together, but I think it’s just something that’s already being looked at from all the different disciplines.”* [Home A]

Other healthcare providers believed PREVENT was a tool to ensure the staff were performing their fall prevention duties, rather than a model of change to prevent fractures in LTC.

> “*It’s worked well just as a tool to let us know if there’s any missing boxes that we’re not checking for fracture prevention. It’s very helpful with just making sure we’re looking at everything and yea, just crossing off any things we can possibly to do help the residents.”* [Home A]

Additionally, several healthcare providers were unsure about the ethical implications of prescribing osteoporosis medications to residents at high risk of fracture. The main barrier to prescribing medications was fear of side effects (i.e., osteonecrosis of the jaw, atypical femur fractures, hypocalcemia). Some providers were also concerned about the ethical implications of prescribing osteoporosis medications to residents who could not consent or had limited knowledge of the medication.

> “*We are worried that, we weren’t overly confident that would lead to prescription. So there was a trigger. We didn’t know if they were going to have informed consent and we didn’t know if our nursing staff knew enough about these medications to provide it*” [Home B].

#### Facilitating factors

Despite some challenges, we also identified several facilitators to our model. Facilitators included easy access to the FRS, clear and succinct educational material catered to each healthcare professional, and an educational module with an accredited program for physicians and nurses. All three homes deemed it beneficial to have the educational module delivered in person. We also received positive feedback on the type of information included in our educational module. In particular, healthcare providers found it helpful to review real-world case studies followed by discussions and suggestions to overcome barriers to implementation (e.g., using hip protectors for residents with incontinence).

> “*A visual to see the walkers and those pieces, I think even the slide with the hip protectors, like how to apply them and which ones were incorrect. I think that’s a nice reminder because I didn’t even know how to position them correctly and I didn’t even have awareness of the FRS. Like I kind of knew like our FRAT* <Falls Risk Assessment Tool> *score and our falls risk, and when I go to my units I know who the falls, the higher risk of falls are but I didn’t, and then I just thought of them like okay, we have hip protectors for them, like what are we doing about it but I didn’t necessarily do a full review of like let’s look at their meds and let’s look at their hips, are they wearing hip protectors, what are they, let’s look at all these other things. I wasn’t sort of looking at that*.” [Home C].

#### Adaptations

The process of developing and implementing models into practice is an iterative process. Through this process, we recognized three adaptations to the PREVENT model including identifying additional characteristics for a successful local champion, developing an automated audit report, and creating tools to facilitate conversations for prescribing osteoporosis medications. It was suggested that the role of the local champion include co-championship with at least two healthcare professionals to help manage the implementation process. Suggested co-championship include one champion be either a registered nurse or a nurse practitioner and the second champion be a registered practical nurse or resident case manager. The benefit of including a resident case manager as a local champion is their established rapport with residents and caregivers and their ability to link residents to other needed services (e.g., referral to a dietitian). Additionally, the local champion must have a basic understanding of medications and have strong collaborations with other healthcare providers including physicians, nurses, dietitians, pharmacists, and physiotherapists. Other adaptations for future practice include working with the electronic medical record company to automate the audit report process, develop conversational toolkits (e.g., between nurses and residents) to aid dialogues about fracture prevention, and develop one-page educational guides for healthcare professionals and residents about the side effects of osteoporosis medications.

### Audit report

We found that the educational module did not improve knowledge uptake of the LTC recommendations among healthcare providers particularly in utilizing the FRS, administrating osteoporosis medications and supplements (calcium and Vitamin D), using hip protectors, and reviewing environmental hazards (mean 0.07, 95% confidence intervals [CI] -0.02 to 0.16). However, our intervention resulted in an absolute increase in the proportion of osteoporosis medications by 6% (Table 4). Home A reported an absolute change of 13%; 21% of residents were on an osteoporosis medication at baseline while post-intervention 34% of residents were on a medication (odds ratio [OR] 0.25, 95% CI 0.12 to 0.48). Similarly, Home B reported an absolute change of 3% (31% at baseline and 34% post-intervention, while Home C had an 8% increase (29% at baseline and 37% post-intervention).

**Table 4:** Aggregated pre-and-post audit report.

|  | **Homes A, B and C** |  |
| --- | --- | --- |
|  | **Pre-intervention** | **Post-intervention** |
| Total number of residents in Home A, B, and C | 739 | 734 |
| Number and percentage of residents at high risk of fracture | 419 (57%) | 430 (59%) |
| Number and percentage of residents at low risk of fracture | 320 (43%) | 304 (41%) |
| Number and percentage of residents who sustained a fragility fracture in the last six months | 20 (3%) | 25 (3%) |
| Number and percentage of residents at high risk of fracture on osteoporosis medications (unspecified amount of time) | 122 (29%) | 150 (20%) |

Note: The number of residents pre-and-post intervention are different because of the “moved-in” (i.e., moved into the LTC home) and “moved-out” (i.e., passed away).

## Discussion

Falls and hip fractures are a major health concern among older adults living in LTC (3,6–8). The purpose of the iCARE study was to determine the feasibility (recruitment rate, facilitators, barriers, adaptations) of implementing PREVENT in LTC homes in Canada. The model includes a multifactorial intervention on diet and supplements, exercise, environmental adaptations, hip protectors, and medication reviews to treat residents at high risk of fracture (9). The standardized model utilizes local champions, educational outreach methods, audit and feedback reports, and implementation intentions to change behaviour. Although we did not meet our recruitment criterion to recruit three homes in two months, our process was affected by COVID-19. Nevertheless, it is possible to recruit LTC homes by accounting for a longer recruitment period (i.e., three homes over five months). We did meet our criterion to recruit a local champion and provide training within our timeframe. Our local champions successfully delivered PREVENT with a fidelity score ranging from moderate to good. We also identified several facilitators, barriers, and adaptations to PREVENT. Benefits of the model include easy access to the FRS, clear and succinct educational material catered to each healthcare professionals, and an educational module with an accredited program for physicians and nurses. We also identified a few challenges such as misperceptions between the differences in fall and fracture prevention strategies, fear of prescribing osteoporosis medications, and time barriers with completing the audit report. Our study did not find a statistically significant increase in knowledge uptake regarding recommendations for preventing falls and fractures; however, there was an increase in the proportion of osteoporosis medication prescriptions post-intervention. The development and implementation of knowledge translation models is an iterative process. This study identified additional changes to PREVENT including co-champions, automated audit report, and tools to facilitate conversations when prescribing osteoporosis medications. Our next step will be to lead a large, pragmatic randomized controlled trial to implement PREVENT in 122 homes across Ontario, Canada.

Effective and sustainable knowledge translation models are essential to encourage the uptake of evidence-based practices (11). The greatest limitation is that there is limited evidence on effective and sustainable models in practice and such models may not be generalizable from one setting to another (16). Compared with community or acute care settings, there has also been limited implementation models for LTC, especially focused on fracture prevention. PREVENT is a scalable educational outreach model that utilizes a multifactorial intervention to reduce falls and fractures among residents in LTC (39). Our study provides preliminary data that suggests PREVENT may be successfully implemented within the LTC setting, particularly when structured to fit the unique practice environment and organizational structure. Most knowledge translation models have been evaluated in non-LTC settings (39). A 2009 Cochrane review reported improvements in care that ranged from 4% to 12% using educational meetings, educational outreach programs, local champions, audit and feedback reports, and computerized reminders (39). When designing PREVENT, we utilized such techniques (e.g., educational meetings, local champion, audit and feedback) as a method to change behaviour (25,27,30,31,39). We observed improvements similar to the Cochrane review ranging from 3% to 13% in the proportion of osteoporosis medications before and after the intervention. Four months was not enough time to implement change, especially in homes with over 200 beds. We saw greater improvements in prescriptions being filled in homes that had a lower rate of medication use. We specifically focused on osteoporosis medications as the highest certainty of evidence to reduce the rate and risk of fractures is using such medications (19,20) and is the easiest outcome to measure. We hypothesize that an increased uptake of osteoporosis medications will translate to lower fracture rates in LTC; however, a large trial is needed to confirm the hypothesis.

Similar models to prevent fractures in LTC include the Bavarian Fall and Fracture Prevention model in Germany that focuses on exercise, documentation of falls, environmental adaptations, medication reviews, vitamin D, hip protectors, and education among healthcare providers (40,41). The Bavarian Fall and Fracture Prevention study is one of the few large studies of an educational outreach program to implement fracture prevention guidelines into LTC (40,41); the authors implemented a multifactorial educational outreach program and after one-year found an 18% reduction in hip fracture in residents from the intervention home (hazard rate ratio 0.82, 95% CI 0.72-0.93) (40,41). While the model was effective, it was a non-randomized, quasi-experimental design that utilized insurance company staff to facilitate and disseminate knowledge on fall and fracture prevention. The sustainability or scalability of this model may not be applicable in the Canada healthcare system. Thus, the next steps will be to conduct a large randomized controlled trial of the adapted PREVENT model in several Canadian LTC homes to determine if the model reduces the rate of hip fractures in residents at high risk of fracture.

Our PREVENT model attempted to leverage the facilitators and limit barriers to implementation as described in other studies. A recent systematic review by our group identified several barriers to implementing guidelines into LTC settings including time constraints and inadequate staffing, cost and lack of resources, knowledge gaps, and lack of teamwork and organizational support (14). Our model addressed such barriers including time constraints, lack of resources, and knowledge gaps by developing a multi-modal approach to embed a validated computer decision support tool and the FRS into already established care pathways. PREVENT also provides an accredited Continuing Medical Education module to physicians and nurses as a method to disseminate evidence-based knowledge. We worked with the home’s management team to identify potential local champions who could help lead the educational module and lead best practices after the research team left; from the literature we identified characteristics for an effective local champion (25,42). Through the iCARE study, we identified additional characteristics to identify a local champion including having co-championship to manage the implementation process and developing conversational toolkits to aid dialogues between healthcare providers and residents on osteoporosis medications. We are also working with a Canadian electronic medical record company to develop an automated audit and feedback report to address the time barrier. We suggest that future knowledge translation and implementation science researchers look to our adapted PREVENT model as there may be important changes to help facilitate the effective translation of evidence into practice.

The iCARE study had several methodological strengths including the recruitment of homes that were geographically diverse and located in communities of varied population sizes; we also included not-for-profit and for-profit homes. Our study implemented a multifactorial intervention that engaged a wide variety of healthcare providers. In addition, our study builds on previous knowledge of implementing models into practice and was designed to leverage facilitators and address common barriers to implementation. To date, there are very few models on chronic disease management especially in LTC (43,44), and so, more studies are needed to identified challenges to implementing knowledge translation models into practice. Nonetheless, our study is not without limitations. Given that our intervention was multifaceted, it was challenging to identify the most significant components (e.g., educational outreach, local champion, audit and feedback report) that would result in effective implementation of PREVENT. Moreover, measures of organizational context such as work culture and leadership style, were not captured in the iCARE study which could have a significant impact on our model’s components. Healthcare sustainability remains a multi-dimension problem, and so it is unclear if local champions or co-championship is sustainable (45). Lastly, for this study, we did not involve residents or their family caregivers during the implementation process. We acknowledge that all residents have complex needs and multiple healthcare issues, and their preferences for care should be incorporated in the decision-making process. Future adaptations for the PREVENT model will be to include end-users on the research team to aid the implementation process to consider how resident’s preferences can be considered. Before we implement the PREVENT model as a randomized controlled trial, we will consult and collaborate with knowledge users on our implementation process.

## Conclusion

PREVENT is a multifactorial educational outreach model to implement education on diet, exercise, environmental adaptations, hip protectors, medications (including calcium and vitamin D) and medication reviews to treat residents at high risk of fracture. The model utilizes educational meetings, local champions, and audit and feedback as methods to promote behaviour change. The model requires ten months to recruit, train, and implement PREVENT, with five of the ten months dedicated toward recruitment of LTC homes. We suggest co-championship when delivering multifactorial models in LTC and developing tools that can be easily embedded into routine practice. Our study did not increase knowledge uptake of the guidelines; however, we did observe an increase in the proportion of osteoporosis medication prescriptions post-intervention. Our next steps will be to consult and collaborate with knowledge users on PREVENT and then lead a large, randomized controlled trial to implement PREVENT to determine if the model reduces the risk and rate of hip fractures in residents living in LTC.

**Figure 1:**
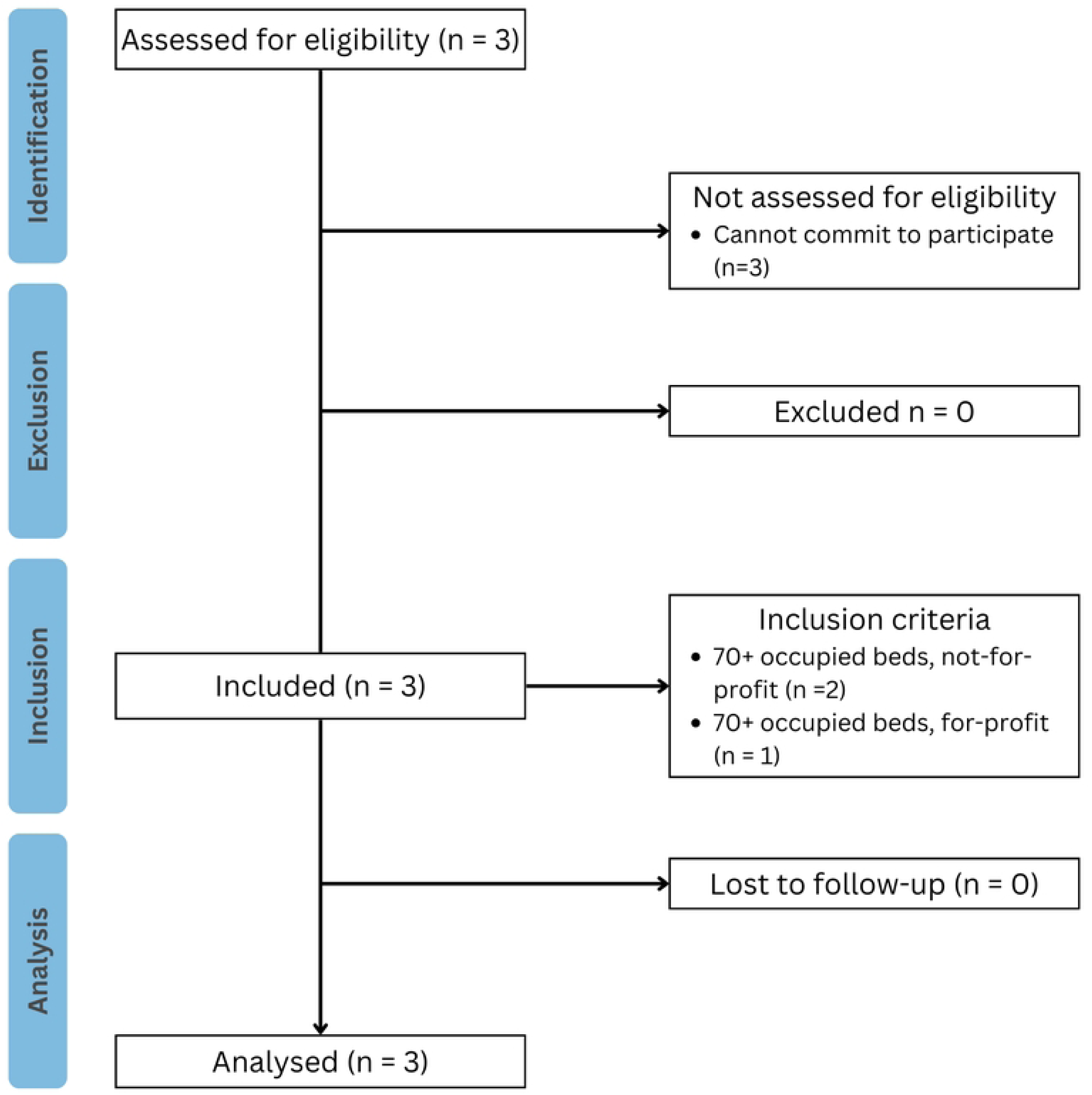
STROBE flow chart. STROBE, Strengthening the Reporting of Observational Studies in Epidemiology.

## Data Availability

Data available upon request from corresponding author

## Ethics approval and consent to participate

In accordance with local regulatory guidelines and standards for human participants’ protection, all methods were carried out in accordance with relevant guidelines and regulations. Our study was reviewed by the Hamilton Integrated Research Ethics Board (HiREB), which is associated with McMaster University. Before enrolling in the study, all participants were briefed on the study and written informed consent was obtained. This review was registered on clinicaltrials.gov (NCT05445336).

## Consent for publication

Not applicable.

## Availability of data and materials

The data generated during the current study are not publicly available due to content that could compromise research participant privacy but are available from the corresponding author on reasonable request for research purposes only.

## List of abbreviations

CI: Confidence Intervals
FRS: Fracture Risk Scale
iCARE: Integrated <u>c</u>ollection of education modules for f<u>a</u>ll and fracture pr<u>e</u>vention
LTC: Long term care
PREVENT: <u>P</u>erson-centred <u>R</u>outine Fracture Pre<u>EVENT</u>ion
OR: Odds Ratio

## Supporting information

S1 File. PREVENT Components

S2 File. Training Manual for Local Champions

S3 File. Fracture & Fall Prevention Guide for Long Term Care S4 Table. Osteoporosis Drugs

S5 Table. STROBE Checklist S6 Table. TIDieR Checklist S7 File. iCARE protocol

